# Machine Learning Models Enhance Prediction of Arrhythmogenic Right Ventricular Cardiomyopathy

**DOI:** 10.1101/2025.06.16.25329706

**Authors:** Kwaku K Quansah, Sean A Murphy, Esther Kwon, Emma Anderson, Crystal Tichnell, Brittney Murray, Sean Gaines, Alessio Gasperetti, Cynthia A James, Hugh Calkins, Richard T Carrick, Chulan Kwon

## Abstract

Arrhythmogenic Right Ventricular Cardiomyopathy (ARVC) is a leading contributor to sudden cardiac death worldwide in young adults, yet its diagnosis remains complex, expensive and time-consuming. Machine-learning (ML) classifiers offer a practical solution by delivering rapid, scalable predictions that can lessen dependence on expert interpretation and speed clinical decision-making. Here, we benchmarked six ML algorithms for ARVC detection using area-under-the-curve (AUC) and accuracy as primary metrics. Gradient Boosted Trees outperformed all other models, achieving a c-statistic of 94.34% after rigorous cross-validation. These results underscore the promise of Gradient Boosted Trees classifier as an effective decision-support tool within the ARVC diagnostic workflow, with potential to streamline evaluation and improve patient outcomes.

## 1. Introduction

Arrhythmogenic Right Ventricular Cardiomyopathy (ARVC) is a hereditary structural heart disease characterized by the progressive replacement of right ventricular myocardium with fibro-fatty tissue.^1,2^ This pathological remodeling results in regional wall motion abnormalities and facilitates occurrence of ventricular arrhythmias, thereby elevating the risk of sudden cardiac death (SCD).^3^ Early identification of ARVC is critical to prevent life-threatening ventricular arrhythmias and SCD, which often occur in young individuals and athletes.^4,5^ The arrhythmogenic substrate in ARVC may be present before overt structural changes appear on imaging, and exertion can precipitate malignant arrhythmias in susceptible individuals.^6–9^ As a result, timely diagnosis allows for the implementation of lifestyle modification such as activity restriction that may limit disease progression, life-saving interventions such as implantable cardioverter-defibrillator (ICD) placement, and family screening.^10–12^ This emphasis on early recognition is reflected in the revised 2010 Task Force Criteria, which sought to standardize diagnostic thresholds by incorporating electrocardiographic, imaging, histological, and genetic parameters.^13,14^ Furthermore, the position statement by the Heart Rhythm Society underscores the importance of early detection and risk stratification to guide preventive therapy.^15^ Longitudinal data from ARVC registries consistently show that delayed diagnosis is associated with increased arrhythmic burden and worse prognosis, highlighting the need for improved screening tools and early intervention pathways.^16–18^

Despite advances in diagnostic criteria, accurately diagnosing ARVC remains a significant clinical challenge.^19,20^ Borderline and gene-elusive presentations often exhibit subtle, variable, or inconclusive findings, leading to high rates of referral for expert re-evaluation.^21–23^ Specialized ARVC centers serve as a referral hubs, yet many referred patients ultimately do not meet diagnostic criteria.^19,24^ This places a substantial burden on the limited number of expert centers and also exposes patients to unnecessary interventions, lifestyle restrictions, and psychological distress resulting from misdiagnosis.^25^ The high volume of false-positive referrals and the potential for clinical mismanagement highlight the need for more efficient and accurate triage strategies.

Machine learning (ML) and deep learning (DL) approaches offer powerful tools to enhance diagnostic precision by identifying complex, nonlinear patterns across multimodal clinical data that may be missed by traditional methods.^26–28^ Significant effort has been devoted to applying DL models to raw electrocardiogram (ECG) and imaging data to improve diagnostic accuracy.^29,30^ For example, Carrick et al. recently developed an ECG-based deep learning (ECG-DL) model that interprets raw ECG waveforms to aid in ARVC diagnosis. While this model achieved a modest but clinically meaningful improvement over expert ECG interpretation (c-statistic 0.87 vs. 0.85), diagnostic performance rose substantially (c-statistic 0.94) when ECG-DL outputs were combined with non-ECG clinical features from the 2010 TFC.^31^ These findings emphasize the added value of multimodal data integration and suggest that future gains in diagnostic accuracy may come more from strategic model design and data fusion than from increasing algorithmic complexity alone.

Therefore, the aim of ths study was to develop, validate, and compare a range of predictive models for ARVC using machine learning algorithms that span from traditional, interpretable approaches to modern, high-complexity techniques. Using a multimodal dataset derived from the Johns Hopkins ARVC Registry, we further compared test characteristics of the top-performing models on hold-out test cohort.

## 2. Methods

### Study Population

For this project, a retrospective cohort was assembled from the Johns Hopkins ARVC Registry to serve as the basis for algorithm training and internal validation.^31^ Patients were included in the cohort if they underwent clinical evaluation for suspected Arrhythmogenic Right Ventricular Cardiomyopathy (ARVC) at Johns Hopkins Hospital (JHH) and had available pre-referral clinical records for review. Inclusion required participation in a standardized second-opinion assessment by a cardiologist with ARVC-specific expertise and a genetic counselor, including review of family history and genetic test results. Diagnostic classification of ARVC was based on the 2010 Task Force Criteria (TFC), requiring patients to meet either two major criteria, one major plus two minor criteria, or four minor criteria from distinct categories. Only patients with available 12-lead ECGs recorded at 500 Hz and stored in the GE Muse system were included, allowing for coding and adjudication of TFC-defined repolarization and depolarization/conduction abnormalities by both the referring cardiologist and an ARVC expert. Patients were excluded if they lacked sufficient clinical documentation, ECG data, or did not undergo comprehensive evaluation at JHH consistent with study protocols.^31^ For each patient, a comprehensive dataset comprising 32 features was constructed by integrating clinical, demographic, and genetic variables relevant to the diagnosis of ARVC.

A composite dataset encompassing all 688 cases was constructed, integrating 30 variables spanning clinical, demographic, and genetic features relevant to ARVC diagnosis. An overview of the data processing and analysis workflow is presented in **Fig. 1**.

**Figure 1.**
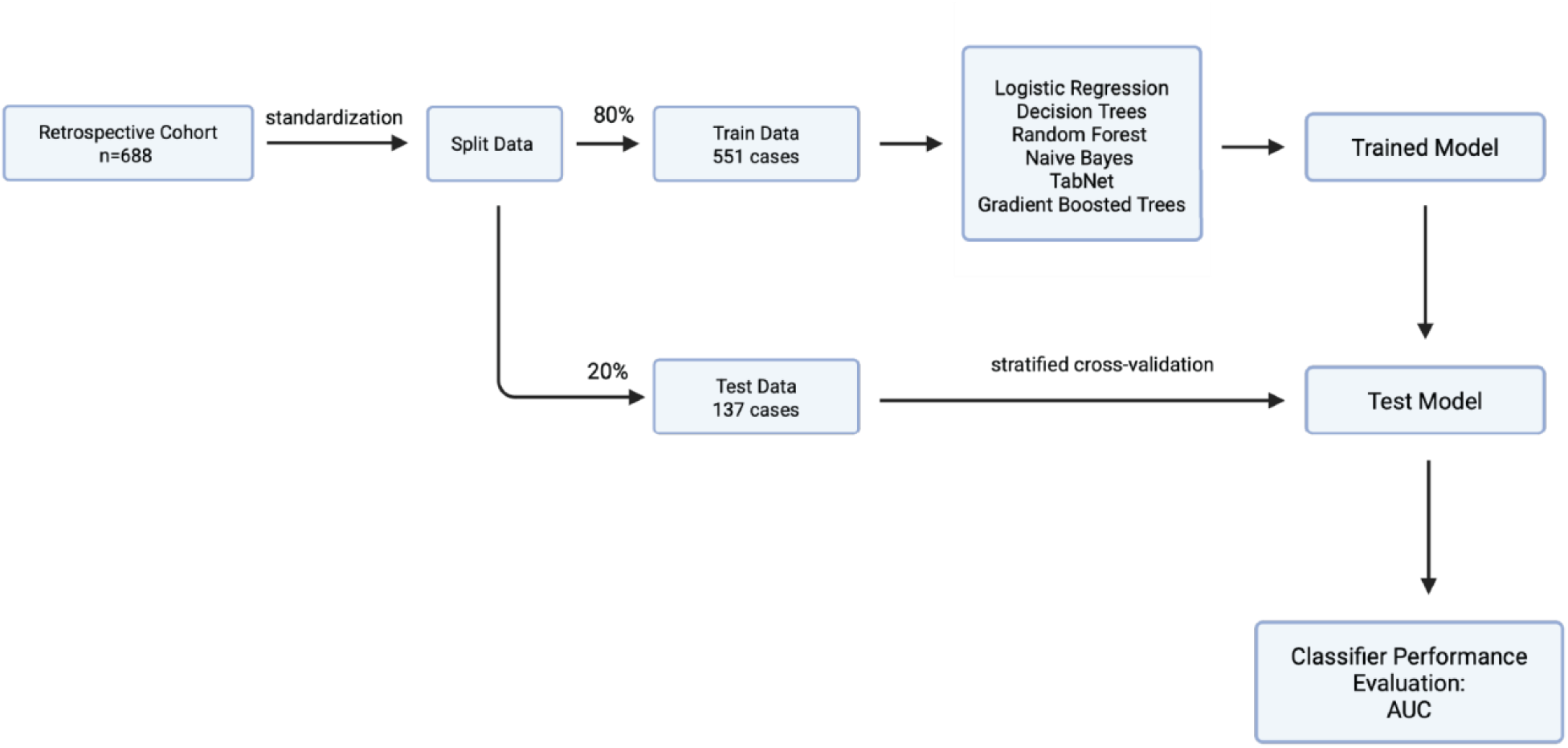
Diagram showing the data split for model training and creation of 8 machine learning models.

### Data Preprocessing

The initial dataset comprised 688 patient records, each containing clinical, demographic, electrocardiographic, imaging, and genetic features extracted from the Johns Hopkins ARVC Registry. These features were derived from diagnostic evaluations, including genetic testing, electrocardiograms (ECG), Holter monitoring, and cardiac magnetic resonance imaging (MRI). Categorical variables such as ‘Ethnicity’ were transformed into numerical input by one-hot encoding.

Approximately 34% of patients had missing values in at least one feature; specific variable missingness is reported in Table 1. For features with missing values, imputation was performed using a k-nearest neighbors (KNN) algorithm with k = 1, enabling preservation of local data structure while maintaining dataset integrity. The final analytic dataset comprised a matrix of 688 patients with 30 complete or imputed feature sets.

**Table 1.** Characteristics of the patient cohort Summary of cohort baseline characteristics. Categorical variables were summarized as frequencies (%) and compared using Chi-square testing. Continuous variables were presented as mean ± standard deviation or median [interquartile range (IQR)], and compared using independent sample Students t-tests or analysis of variance (ANOVA), respectively. SCD_1deg_relative_under35 = Sudden Cardiac Death in a first-degree relative under 35; TWI = T wave Inversion; PVC = Premature Ventricular Contractions; LGE = Late Gandolinium Enhancement; PKP2 = Plakophilin 2; DSP = Desmoplakin; DSG2 = Desmoglein-2; DSC2 = Desmocollin-2; CRBBB = Complete Right Bundle Brunch Block; CLBBB = Complete Left Bundle Brunch Block; TAD = Terminal Activation Duration; LVEF = Left Ventricular Ejection Fraction; RVEF = Right Ventricular Ejection Fraction.

| Characteristic | All Cases | % Missing | ARVC cases | Non-ARVC cases | P-value |
| --- | --- | --- | --- | --- | --- |
| Sample size | 688 |  | 329(47.8%) | 359(52.2%) | chi-squared test for categorical data |
| Age | 40.2±15.1 | 0 | 39.41 ± 14.69 | 40.84 ± 15.48 | 9.50E-04 |
| Sex |  | 0 |  |  | 0.0013 |
| Male | 394(57.3%) |  | 167(50.76%) | 227(63.23%) |  |
| Female | 294(42.7%) |  | 162(49.24%) | 132(36.77%) |  |
| Ethnicity |  | 0 |  |  | 0.8456 |
| White | 576(83.7%) |  | 274(83.28%) | 302(84.12%) |  |
| Non-white | 112(16.3%) |  | 55(16.72%) | 57(15.88%) |  |
| SCD_1deg_relative_under35 | 118(17.15%) | 0.15 | 73(21.6%) | 45(12.4%) | 0.0012 |
| Pathogenic variants in *PKP2* | 152(28.35%) | 0 | 124(37.7%) | 28(7.79%) | 0 |
| Pathogenic variants in *DSP* | 49(7.12%) | 0 | 33(10%) | 16(4.4%) | 0.0071 |
| Pathogenic variants in *DSG2* | 31(4.5%) | 0 | 28(8.5%) | 3(0.8%) | 0 |
| Pathogenic variants in *DSC2* | 13(1.8%) | 0 | 11(3.3%) | 2(0.5%) | 0.0163 |
| Mutation in other Gene | 35(5%) | 0 | 22(6.7%) | 13(3.62%) | 0.0981 |
| TWI_V1 | 391(56.91%) | 0.15 | 265(80.55%) | 126(35.20%) | 0 |
| TWI_V2 | 295(42.94%) | 0.15 | 238(72.64%) | 56(15.64%) | 0 |
| TWI_V3 | 288(41.92%) | 0.15 | 248(75.38%) | 40(11.17%) | 0 |
| TWI_V4 | 205(29.84%) | 0.15 | 183(55.62%) | 22(6.15%) | 0 |
| TWI_V5 | 126(18.34%) | 0.15 | 115(34.95%) | 11(3.07%) | 0 |
| TWI_V6 | 77(11.21%) | 0.15 | 72(21.88%) | 5(1.40%) | 0 |
| TWI_II | 16(2.33%) | 0.15 | 15(4.56%) | 1(0.28%) | 0.0005 |
| TWI_III | 35(5.09%) | 0.15 | 25(7.60%) | 10(2.79%) | 0.0072 |
| TWI_aVF | 27(3.94%) | 0.29 | 20(6.10%) | 7(1.96%) | 0.0096 |
| CRBBB | 49(7.13%) | 0.15 | 23(6.99%) | 26(7.26%) | 1 |
| CLBBB | 2(0.29%) | 0.29 | 0(0.00%) | 2(0.56%) | 0.5151 |
| LGE | 170(34.00%) | 27.33 | 133(56.12%) | 37(14.07%) | 0 |
| TAD | 34(4.95%) | 0.15 | 24(7.29%) | 10(2.79%) | 0.011 |
| Ventricular_Rate | 65.15 ± 14.36 | 0 | 64.29 ± 15.70 | 65.95 ± 12.99 | 1.33E-01 |
| PR_Interval | 158.93 ± 39.13 | 0 | 159.42 ± 43.92 | 158.48 ± 34.22 | 7.54E-01 |
| QRS_Duration |  | 0 |  |  | 9.63E-02 |
| QT_Corrected | 426.04 ± 33.92 | 0 | 431.39 ± 34.96 | 421.13 ± 32.22 | 7.38E-05 |
| R_Axis | 38.53 ± 53.34 | 0 | 41.80 ± 61.11 | 35.54 ± 44.94 | 1.29E-01 |
| LVEF | 56.66 ± 9.36 | 12.79 | 55.77 ± 10.14 | 57.48 ± 8.53 | 1.15E-01 |
| PVC | 3739.73 ± 7033.90 | 15.55 | 4386.20 ± 6567.72 | 3147.29 ± 7395.10 | 2.38E-02 |
| RVEF | 45.28 ± 10.86 | 31.4 | 41.12 ± 10.74 | 49.10 ± 9.49 | 2.26E-16 |

To assess linear correlation among features, we conducted a Pearson correlation analysis across variables. Most features exhibited low-to-moderate correlation, with the highest observed correlation being 0.78 between certain ECG lead variables **(Table1)**. These findings supported the inclusion of all features in the modeling pipeline without redundancy reduction.

### Outcomes

The primary outcome for all analyses was fulfillment of the 2010 modified task force criteria for diagnosis of ARVC after comprehensive clinical assessment at the Johns Hopkins ARVC center.

### Machine Learning Models

Six distinct machine learning algorithms were employed, representing a range of model categories. These included Decision Trees and their ensemble methods—Gradient Boosted Trees and Random Forest—which are effective in capturing nonlinear relationships and complex feature interactions. Linear classifier Logistic Regression was due to its interpretability and efficiency. A probabilistic classifier, Gaussian Naive Bayes, was included for its capacity to handle high-dimensional, sparse data and class imbalance. Additionally, a deep learning–based model, TabNet, was implemented to explore attention-driven neural network performance on structured, tabular data.

All model development and analysis were conducted in Python version 3.10, using a suite of open-source libraries tailored for machine learning applications. We used scikit-learn (v1.2.2) for classical machine learning algorithms and model evaluation utilities, and PyTorch (v1.13.1) for neural network modeling. The TabNet model was implemented using the pytorch-tabnet library (v3.1.1). Additional libraries used for data processing and visualization included NumPy (v1.24.2) and pandas (v1.5.3). To optimize model performance, hyperparameters for all algorithms except Naive Bayes and Logistic Regression were systematically tuned using GridSearchCV, a grid-based search algorithm from scikit-learn.

### Model Testing and Analysis

Model evaluation was performed using an 80/20 train-test split, with 80% of the data allocated for training and cross-validation, and the remaining 20% reserved for independent hold-out testing. Categorical variables were summarized as frequencies (%) and compared using Chi-square testing. Continuous variables were presented as mean ± standard deviation or median [interquartile range (IQR)], and compared using independent sample Students t-tests or analysis of variance (ANOVA), respectively. Model discrimination was assessed by calculating the area under the receiver operating characteristic curve (AUROC) using the roc_auc_score function from the scikit-learn library. The classification task was framed as a binary outcome, where the model output represented the predicted probability of a patient meeting diagnostic criteria for ARVC. Feature importance for Gradient Boosting classifier was quantified using Gini importance (also known as mean decrease in impurity), as implemented in the scikit-learn framework, to identify variables contributing most significantly to model predictions.

Continuous probability outputs from each model were dichotomized into ARVC-positive or ARVC-negative classifications using a prediction threshold of 0.5. Probabilities ≥0.5 were considered indicative of ARVC, while those <0.5 were classified as non-ARVC. This standard cutoff was used to evaluate model performance.

Confusion matrices were generated for each model, from which sensitivity, specificity, and overall accuracy were calculated. Sensitivity was defined as the proportion of true positives among individuals with confirmed ARVC, and specificity as the proportion of true negatives among individuals without the disease. Overall diagnostic accuracy was defined as the proportion of all correctly classified cases. Exact binomial confidence intervals (95% CI) were computed for sensitivity and specificity.

## 3. Results

### Cohort and Feature Characteristics

As previously described, the study cohort included 688 individuals suspected of having ARVC who were evaluated at the Johns Hopkins ARVC Center between January 2011 and September 2019.^31^ Of these, 358 patients (52%) fulfilled ARVC task-force criteria after comprehensive assessment. Mean age was 39 years, 49% were female and 83% were white. 525 patients (76%) underwent genetic testing, with pathogenic or likely pathogenic variant identified in 357 patients (68%). On imaging assessment, mean ejection fraction for RV and LV were 41.1% and 55.8% respectively **(Table 1)**. For the remaining 330 patients who did not fulfill ARVC task force criteria, alternative diagnoses after comprehensive assessment included vasovagal syncope, idiopathic ventricular tachycardia, idiopathic pvc, athlete’s heart, ventricular fibrillation from ischemia, cardiac sarcoidosis, orthostatic hypotension, myocarditis, tricuspid regurgitation, sinus node dysfunction, atrial fibrillation with aberrancy, lbbb-induced cardiomyopathy, and pectus-induced dyspnea.

To guide feature selection, Pearson correlation analysis was conducted to assess linear relationships among all variables. Most features exhibited low to moderate correlations, with the highest observed value being 0.78 between specific ECG lead variables (**Fig. 2**). Accordingly, all features were retained in the modeling pipeline, as no evidence of excessive collinearity or feature redundancy was identified.

**Figure 2.**
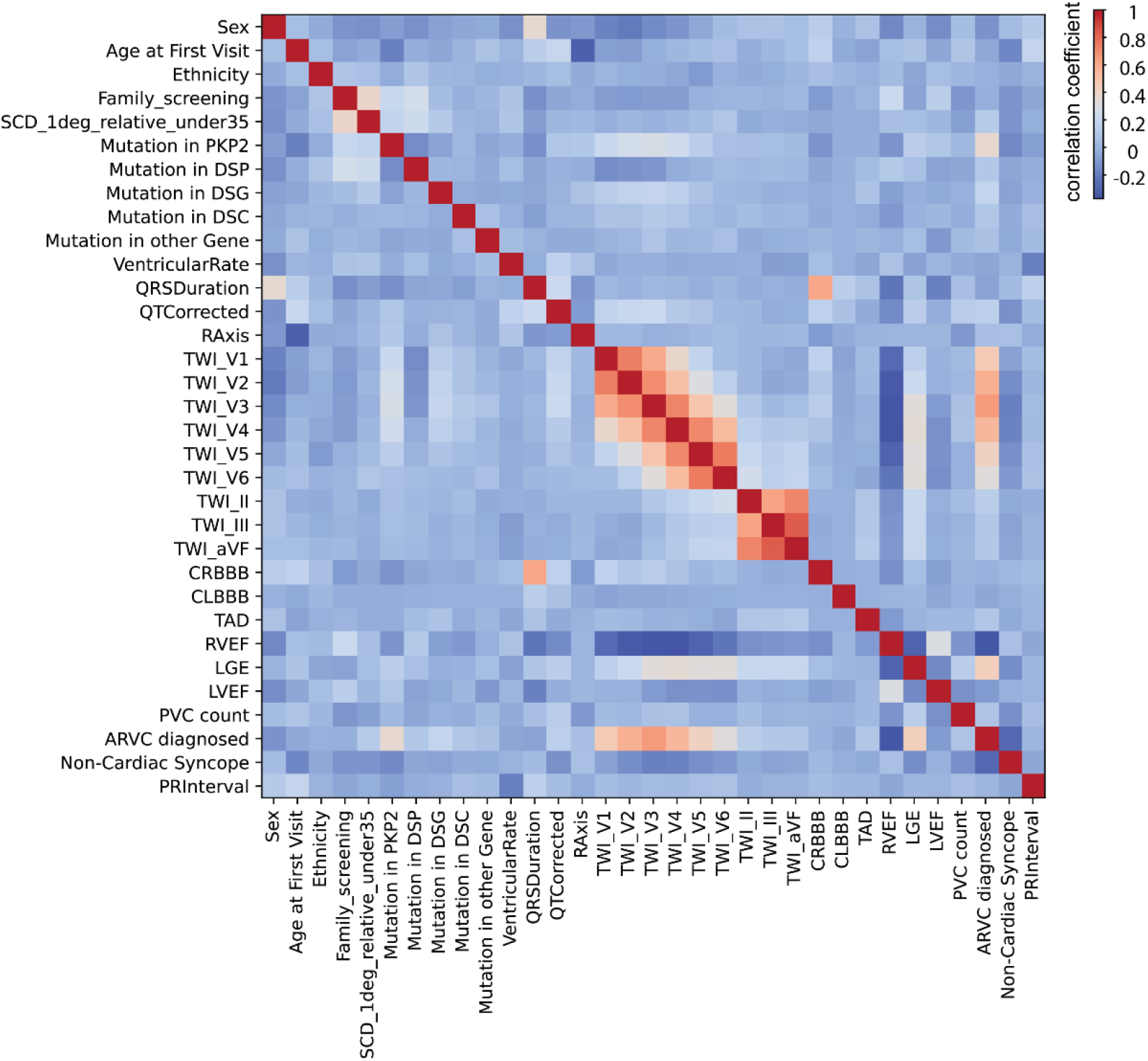
Pearson correlation heatmap with scale bar ranges from red negative correlation to blue positive correlation. Abbreviations are available in the appendix.

### Performance of Predictive Models and Feature Importance

A broad range of ML models were selected including a generalized linear model, DL, and tree models. All six predictive models (Random Forest, Decision Trees, TabNet, Gradient Boosted Trees, Logistic Regression, and Gaussian Naïve Bayes) demonstrated strong discriminatory capacity in the hold-out testing cohort, with mean AUC values exceeding 0.90 **(Supplementary Fig.1)**. Gradient Boosted Trees achieved the highest AUC at 0.943, closely followed by Random Forest (0.938), indicating that ensemble-based methods and regularized linear models are better at capturing feature patterns associated with ARVC diagnosis **(Fig.3**).

**Figure 3.**
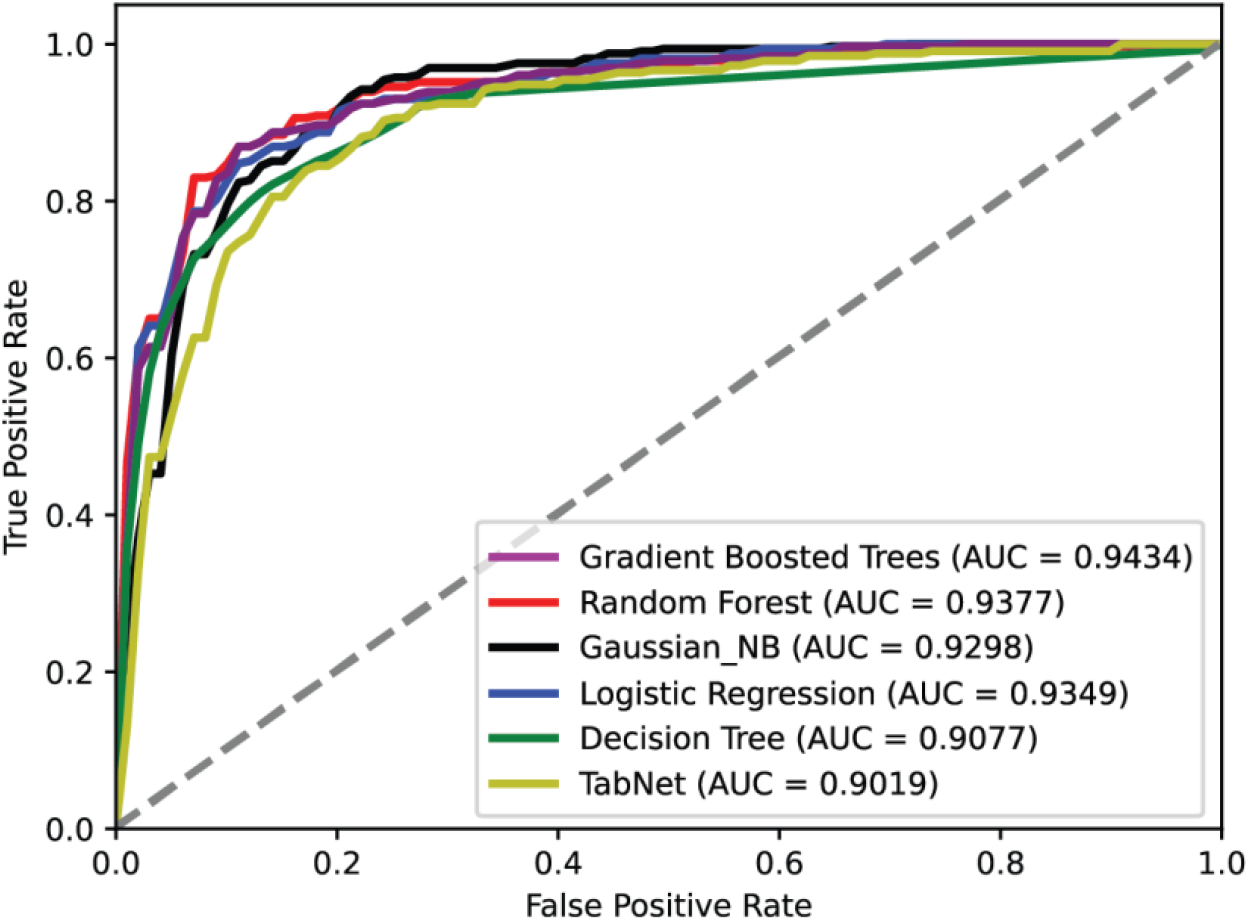
Comparison of ROC of all 8 machine learning models.

Logistic Regression also achieved high AUC of 0.935, suggesting that a linear approach can perform comparably to more complex models. Gaussian Naïve Bayes, a probabilistic model, achieved a slightly lower AUC of 0.929 but still demonstrated strong classification capability.

In terms of classification metrics, Random Forest and Gradient Boosted Trees performed well, with sensitivities of 86.4% and 84.8%, respectively. Notably, Gaussian Naïve Bayes exhibited the highest specificity at 90.1% (95% CI: 81.0%-95.1%), suggesting superior performance in correctly identifying true negatives, though this came at the cost of lower sensitivity (0.758).

Overall accuracy across models was consistently high, ranging from 82.5% to 86.1%. Random Forest had the highest accuracy at 86.1% with Gradient Boosted Trees following closely at 84.7%. All models achieved statistically significant results with p-values < 0.001, reinforcing the robustness of their predictive performance **(Table 2)**.

**Table 2.**
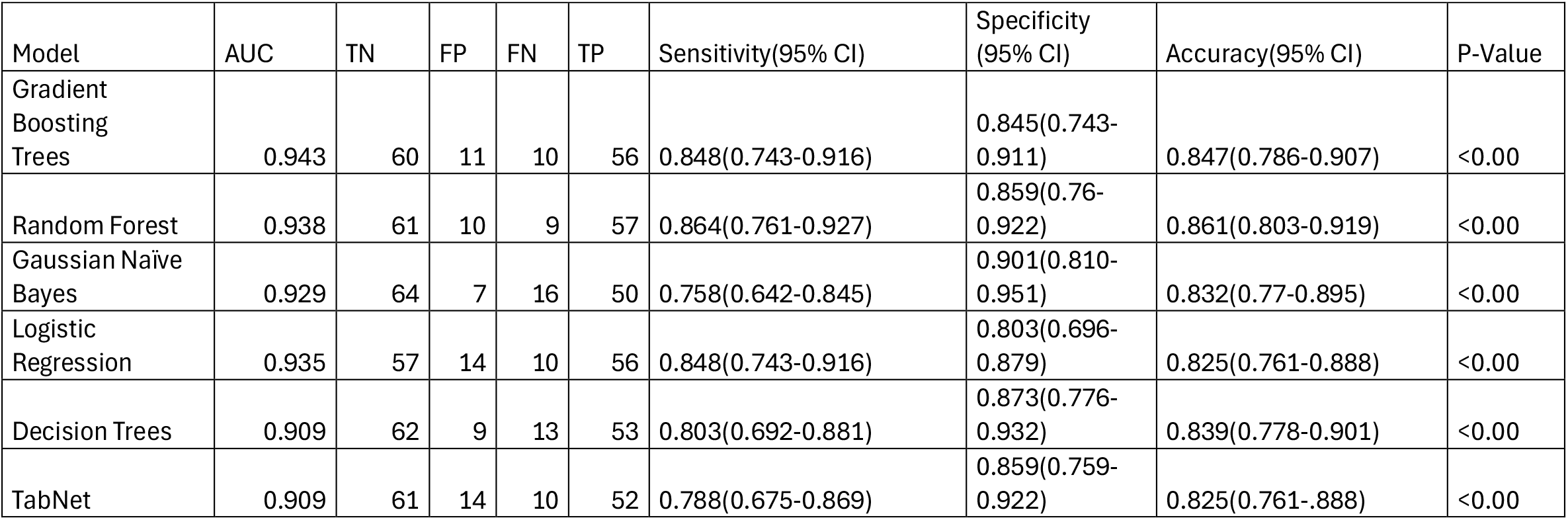
Test characteristics of top ARVC prediction models on hold-out test cohort TN= True Negative; FP= False Positive; FN= False Negative; TP= True Positive; AUC= Area Under the Curve

### Feature Importance by Best Performing Model

The performed saliency analysis for the best-performing Gradient Boosting Trees Model using Gini importance. The model T wave inversion in lead V2 as the most influential features for classifying ARVC, highlighting the strong predictive value of ECG-derived abnormalities **(Fig. 4)**. Structural imaging markers such as right ventricular ejection fraction (RVEF) and late gadolinium enhancement (LGE) also contributed meaningfully, though to a lesser extent. Presence of genetic variants, along with clinical variables like PR interval and age at first visit, showed comparatively lower importance. Overall, the model emphasizes that T wave inversions and arrhythmic burden carry the highest discriminatory power, consistent with their known early presence in ARVC pathogenesis.

**Figure 4.**
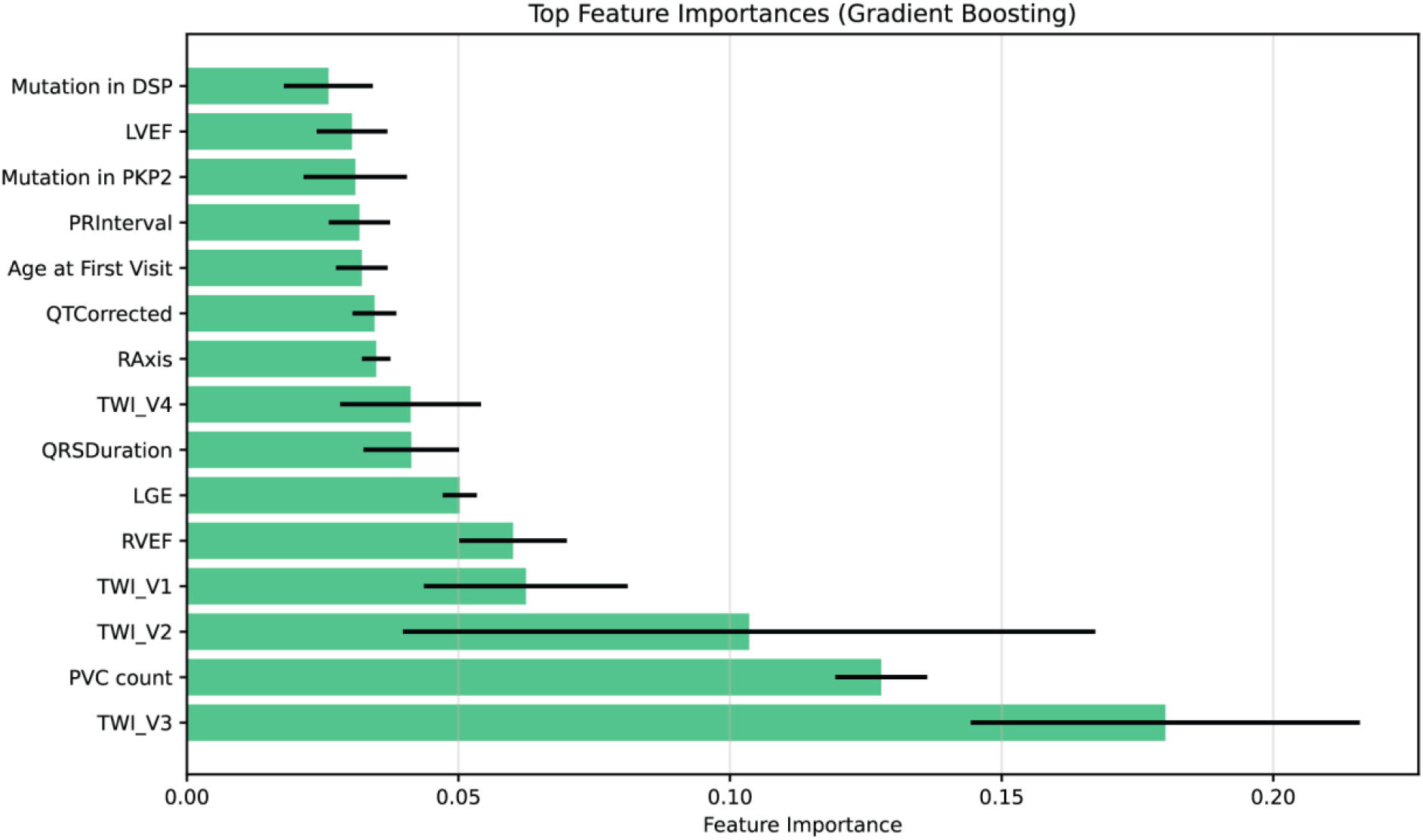
Feature Importance by Gradient Boosted Trees. Abbreviations are available in the appendix.

## 4. Discussion

In this study, we examined a range of alternative machine learning algorithms for diagnosis of Arrhythmogenic Right Ventricular Cardiomyopathy (ARVC), leveraging a multimodal dataset derived from a large referral cohort. Of the examined algorithms, a Gradient Boosting Trees classifier achieved the highest overall performance with an AUC of 0.943 and outperformed both the traditional linear model and more complex deep learning approaches. These results highlight the potential of ML-based decision support tools to improve diagnostic precision for ARVC, a disease where early identification is critical yet often elusive in standard clinical practice.

### Comparison of Alternative ML algorithms

Notably, Logistic regression performed comparably to ensemble methods, achieving AUC exceeding 0.93 while offering greater interpretability. This suggests that, in well-structured datasets, even relatively simple models can achieve clinically actionable performance. In contrast, the TabNet deep learning model underperformed despite its capacity to model attention across structured data, possibly due to limited sample size and the absence of temporally rich or high-dimensional input modalities. These results support the notion that model architecture should be carefully matched to the structure of available data and that increased complexity does not always yield superior performance. Overall, our findings support the use of ML models as adjunctive tools for streamlining ARVC diagnosis, particularly in settings where expert interpretation is limited or delayed.

These findings suggest that simpler, interpretable models may retain considerable diagnostic value while offering the additional benefit of transparency in clinical settings.

In contrast, Decision Trees and TabNet models showed relatively lower performance, with AUCs of 0.910 and 0.900, respectively. While still above the threshold typically considered acceptable for medical classification tasks, their performance lagged behind ensemble methods and the linear model. Overall, these results underscore the importance of model selection in diagnostic tool development for ARVC, with ensemble and regularized linear models emerging as the most effective approaches in this study.

### Model Explainability

A key insight from our feature importance analysis was the prominent role of electrocardiographic markers, specifically, T wave inversion in precordial leads V2 and V3, and ventricular ectopy burden as measured by Holter monitoring. These findings are consistent with known disease mechanisms in ARVC and suggest that surface ECG features, which are both low-cost and readily available, may contain disproportionately high predictive value.^32^ Structural imaging parameters, such as right ventricular ejection fraction and late gadolinium enhancement, contributed to a lesser extent, suggesting that electrical disturbances precede overt structural remodeling in many cases. Interestingly, genetic variants in *PKP2* and *DSP* displayed lower relative importance, which may reflect the variable penetrance of variants in these genesand the presence of both at-risk family members and gene-elusive forms of ARVC in the dataset.

### Clinical Implications

Future work should prioritize external validation of these models in multi-institutional datasets, particularly those with greater ethnic, geographic, and clinical diversity. In addition, integrating unstructured data, including raw ECG waveforms, cardiac imaging, and free-text clinical notes, may enable development of hybrid models capable of capturing temporal dynamics and subtle phenotypic nuances. Prospective evaluation in real-world clinical workflows is also critical to assess whether ML-based tools improve diagnostic efficiency, reduce unnecessary referrals, and optimize patient outcomes. Furthermore, longitudinal models that incorporate time-series data may enhance risk stratification and predict disease progression in individuals with indeterminate or borderline features. Finally, as molecular and genomic testing become more integrated into routine cardiology care, future models should explore the utility of polygenic risk scores and transcriptomic signatures in improving classification performance, particularly in genetically elusive ARVC subtypes. Collectively, these efforts will support the safe and effective translation of ML approaches into clinical practice for heritable arrhythmia syndromes.

### Limitations

This study has several limitations that warrant consideration. First, the data were sourced from a single high-volume referral center, which may limit external validity. The cohort likely reflects a more diagnostically challenging subset of patients, and findings may not generalize to broader, community-based populations.

Furthermore, diagnoses were adjudicated using the 2010 Revised Task Force Criteria, which may not fully capture evolving clinical phenotypes, particularly in early-stage or left-dominant presentations. Finally, while feature importance metrics offer interpretability, they do not establish causality and may be affected by feature collinearity or model-specific biases.

## 5. Conclusion

In summary, this study demonstrates the strong potential of machine learning—particularly Gradient Boosted Trees—in enhancing the diagnostic evaluation of Arrhythmogenic Right Ventricular Cardiomyopathy (ARVC). By benchmarking six distinct algorithms on a richly phenotyped, multimodal dataset from a specialized ARVC registry, we found that ensemble and linear models outperformed both simple classifiers and deep learning approaches in terms of diagnostic accuracy, sensitivity, and specificity. Gradient Boosted Trees achieved the highest overall performance, while interpretability and feature relevance analyses reaffirmed the diagnostic importance of ECG-derived markers such as T wave inversion. These findings suggest that ML-based tools can serve as effective adjuncts in clinical triage and diagnosis, potentially reducing delays, unnecessary referrals, and resource burden on expert centers. Future work should focus on external validation, integration of unstructured and longitudinal data, and prospective deployment in clinical settings to fully realize the benefits of ML-guided decision support in ARVC and related heritable cardiomyopathies.

## Data availability

Patient information was not able to be sufficiently deidentified and is protected. Therefore, is not available.

## Code availability

Code is available on Github at https://github.com/Kwaku-quansah/ARVC_ML.git

## Ethics statement

All work is approved by the Johns Hopkins Hospital IRB.

## Author contribution

K.K.Q, S.A.M and R.C conceptualized the work. K.K.Q and S.A.M developed the methodology. R.C, C.J, B.M, C.T, S.G, A.G and H.C were involved in data curation. C.T, B.M, C.J and H.C worked on required IRB protocols. K.K.Q, S.A.M, E.K, E.A trained, developed and tested the models. K.K.Q, S.A.M and E.K wrote the original draft. K.K.Q, S.A.M, E.K and E.A generated the figures. K.K.Q, C.K, R.C, H.C revised the manuscript.

C.K and R.C supervised the project. C.K provided funding for the project.

---

**Supp. Figure S1.**
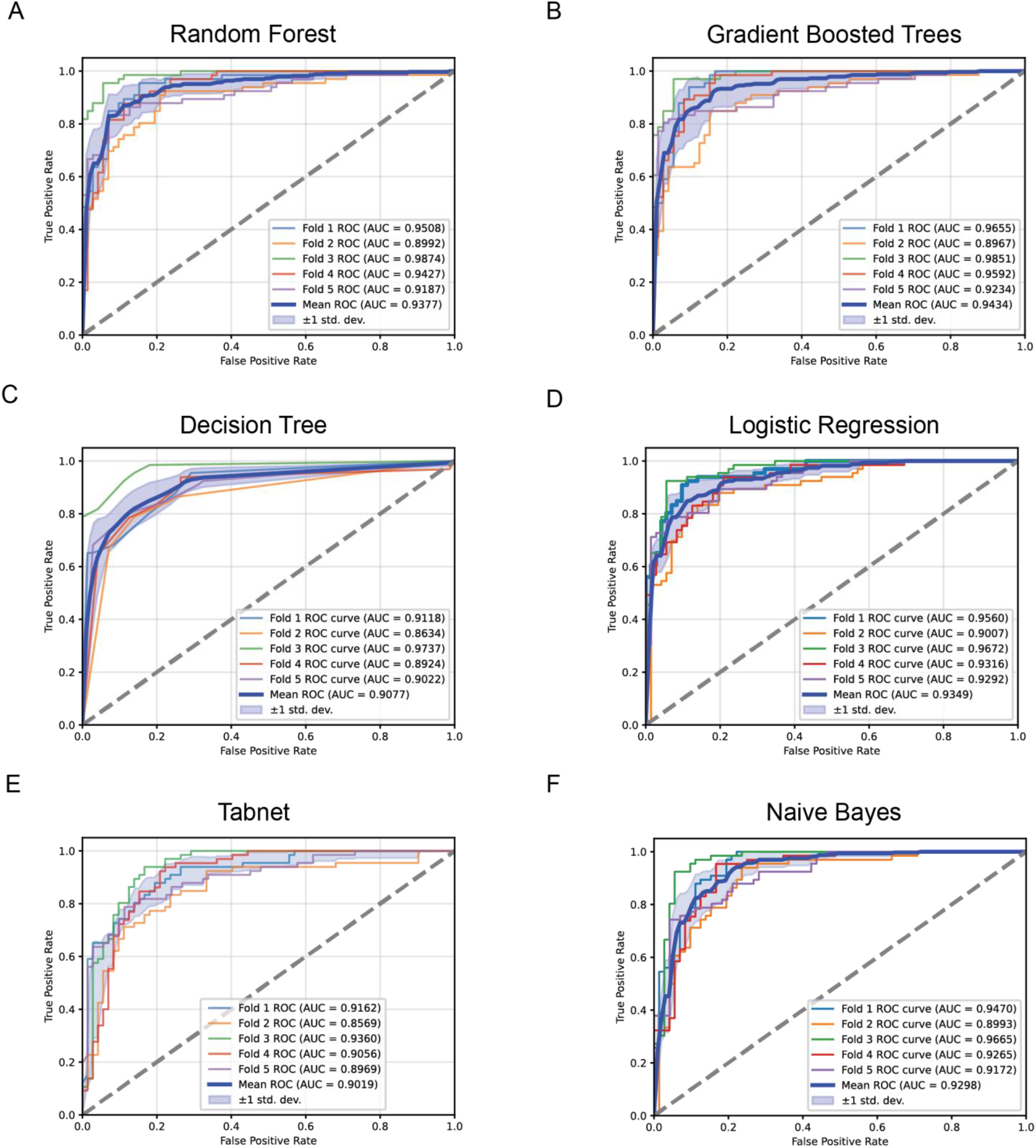
ROC of each trained model A. Random Forest B. Gradient Boosted Trees C. Decision Tree D. Logistic Regerssion E. Tabnet F. Naïve Bayes.

